# Anterior Segment Parameters on Optical Coherence Tomography in Healthy Children

**DOI:** 10.1101/2022.01.31.22270054

**Authors:** Sana Nadeem

## Abstract

**Purpose:** To assess the anterior segment parameters in healthy children, using high resolution spectral domain optical coherence tomography (SD-OCT).

**Design:** Prospective, observational study

**Methods:** 108 eyes of 54 healthy children ≤16 years were included after a thorough eye examination. Significant refractive error (>±5 DS), history of previous glaucoma, ocular pathology, intraocular surgery, trauma or systemic ailments were excluded. The anterior segment analysis was done by Optopol Revo 80^®^ high resolution SD-OCT. Central corneal thickness (CCT), Angle opening distance (AOD), Anterior chamber (AC) angle, Trabecular iris space area (TISA), Trabecular iris angle (TIA), iris thickness at 2 mm, internal AC diameter and lens vault were evaluated.

**Results:** Mean age was 11.38±3.18 years with a female predominance; 29 (53.7%). Mean IOP was 13.63±2.90 mmHg, mean axial length was 22.82±0.89 mm, mean spherical equivalent was -0.14±1.28 D, mean CCT was 532.6±46.09mm and mean CDR was 0.31±0.19. The mean internal AC diameter was 11609.15±2354 µm. The nasal AC angle was 53.54±11.82° and significantly wider than the temporal AC angle; 50.37±10.26° [p=0.008]. The nasal AOD500 was 0.9±0.33 mm and not significantly greater than the temporal AOD500, 0.85±0.28 mm [p=0.09]. The nasal AOD750 was 1.21±0.45 mm, significantly greater than the temporal AOD750, 1.06±0.32 mm [p=0.001]. The nasal TIA was 41.75°; not significantly greater than the temporal TIA, 40.24° [p=0.246]. The nasal TISA500 was 0.33±0.15 mm^2^; not significantly wider than the temporal TISA500, 0.31±0.10 mm^2^ [p=0.056]. The nasal TISA750 was 0. 59±0.22 mm^2^; not significantly wider than the temporal TISA750, 0.56±0.21 mm^2^ [p=0.14]. The nasal iris thickness at 2 mm from pupil was 483.54±129.56 µm; not significantly different from the temporal iris thickness, 505.8±138.85 µm [p=0.08]. The mean lens vault was - 519.58±359µm.

**Conclusions:** Our study data depicts the normal anterior segment parameters in healthy children. This will be useful in diagnosing and managing ocular pediatric pathology.

**Key Messages:**

- Anterior segment OCT is still an emerging technique especially in pediatric eye assessment
- Anterior segment evaluation via OCT has not been reported in our pediatric population
- Our study describes a detailed assessment of anterior segment in pediatric eyes
- Reporting results of normative data helps in diagnosing ocular pathology

## 1. INTRODUCTION

Optical coherence tomography (OCT) is a non-invasive, rapid imaging modality which utilizes infrared light reflected by the ocular structures to create high resolution, cross sectional, three-dimensional images for the assessment of anterior and posterior segment structures [1]. Anterior segment OCT (AS-OCT) is an imaging technique which is rapidly emerging and now becoming a key element in anterior segment evaluation, creating a high impact on clinical practice. It provides a detailed optical section for imaging of the anterior segment, especially the cornea, anterior chamber angle, its structures, iris root, angle recess, anterior ciliary body, scleral spur, Schlemm’s canal, trabeculum, the sclera, iris, and lens [2-4]. It can also be used to measure corneal thickness, anterior and posterior chamber depths, anterior chamber diameter, chamber angle configuration and even iris and lens thicknesses. Mostly, OCT is done on the adult population and there is only limited literature available on pediatric eyes [5, 6]. Currently, AS-OCT is not routinely employed in pediatric ocular evaluation, especially in our country. This is partly due to the fact that some cooperation is required for this study and very small children may not be suitable subjects [5, 6]. Also, in a developing country the availability of OCT is limited to a few specialized centers.

The rationale of our study was to employ AS-OCT in the evaluation of normal children of the Pakistani population and to establish a normative database for anterior segment structures. Studying of normal eyes will help in distinguishing them from pathological cases.

## 2. MATERIALS AND METHODS

A total of 108 eyes of 54 cooperative, healthy children and teenagers between the ages of 3 to16 years presenting to the author either in the out-patient department or in-patient department of Ophthalmology, Fauji Foundation Hospital, from 1^st^ September, 2020 till 22^nd^ February 2021, were included in this observational study, if upon examination, they were found to be ‘*clinically normal’*. A thorough eye examination with visual acuity, refraction (cycloplegic retinoscopy or automated), intraocular pressure estimation by applanation tonometry, axial length estimation by A-scan (Quantel Medical^®^ Axis-II France), anterior and posterior segment examination by slit lamp was done prior to recruiting the children for OCT. Intraocular pressure (IOP) higher than 21 mmHg was excluded, along with any kind of ocular pathology or history of ocular trauma, non-cooperative children, those with systemic diseases and suspicious looking discs. In cases of large cups, glaucoma was excluded by perimetry (Humphrey Field Analyzer Model 720 Carl Zeiss^®^ Meditec Dublin CA, USA) and OCT, before categorizing the child as normal. A spherical equivalent of more than ± 5.00 D was excluded as well.

> ***Permission from the ‘Foundation University Ethical review committee’ [FF/FUMC/215-41 Phy/20] was taken previously, which is in concordance with the declaration of Helsinki. Parental informed consent and patient consent to participate in the study was taken prior to evaluation***.
>
> The Optopol Revo 80^®^ is a high resolution spectral domain (SD-OCT) with a wavelength of 840 nm, a transverse resolution of 12 µm and an axial resolution of 5 µm. It provides 80,000 A scans per second, with a scan depth of 2.4 mm. It provides 3-Dimensional, radial, B-scan, raster, and cross scans. For standard examinations no additional lens is needed. Additional adapter provided with the device allows us to make wide scans of anterior segment.

The procedure was explained to the parents and child and the forehead and chinrest of the Optopol Revo 80^®^ high resolution SD-OCT machine was adjusted for each child. They were assured that it was a non-contact imaging device and the examination was painless and without a topical anesthetic. The child was asked to look into the imaging aperture and focus on the center of the internal fixation target (green cross) and blink freely during the examination. Central corneal thickness was measured first on anterior radial scans, without the adapter provided with the device. The adapter was then mounted in place over the imaging aperture of the OCT machine and the child again reassured. Anterior segment wide scan was taken and anterior radial scans were then taken of each eye separately. Imaging of the angle structures was done with the adapter by manually moving the cursor away from the center and capturing the required images, both nasally and temporally. Good quality images were saved and measurements were performed later. Our OCT machine currently does not have a biometry attachment, so certain measurements like anterior chamber depth and lens thickness were not assessed. Both eyes were taken into consideration in our study. Figure 1 depicts normal anterior segment structures as seen on the OCT.

**Figure 1:**
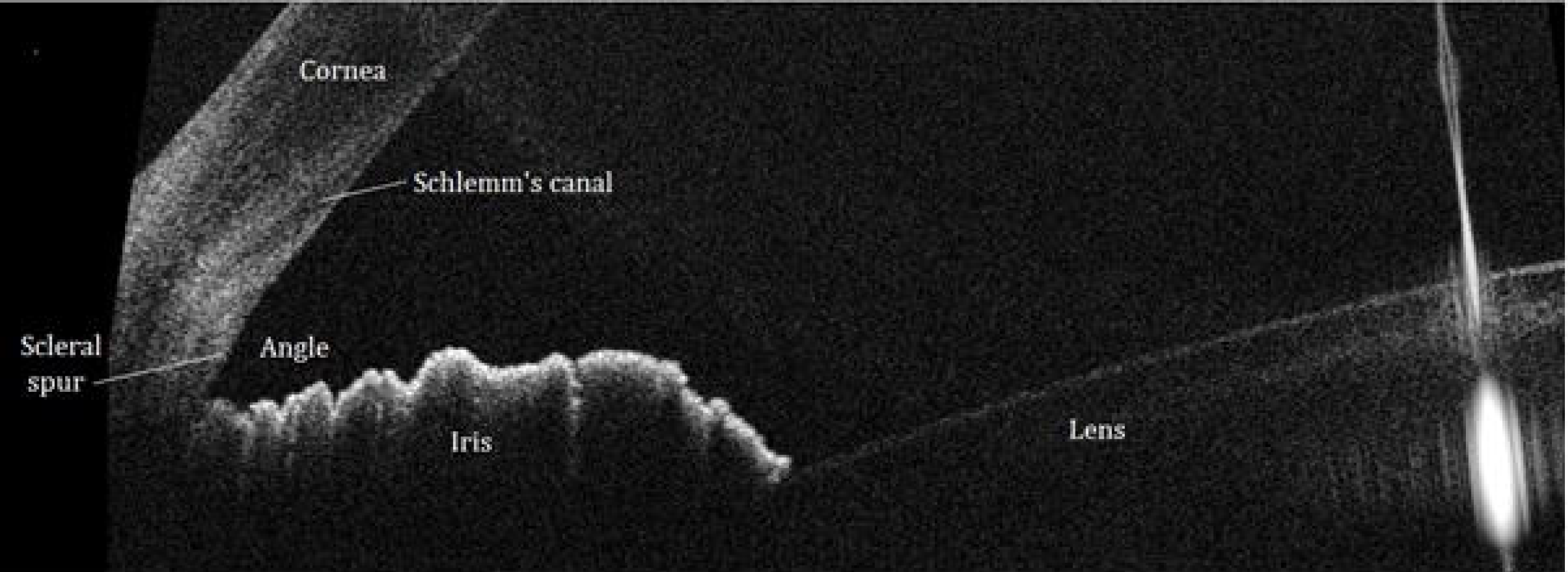
Normal Anterior Chamber structures

### 2.1. Measurements

#### 2.1.1. Central corneal thickness (CCT)

was automatically calculated in mm by the machine as the central distance between the epithelium and endothelium. [Figure 2A]

**Figure 2:**
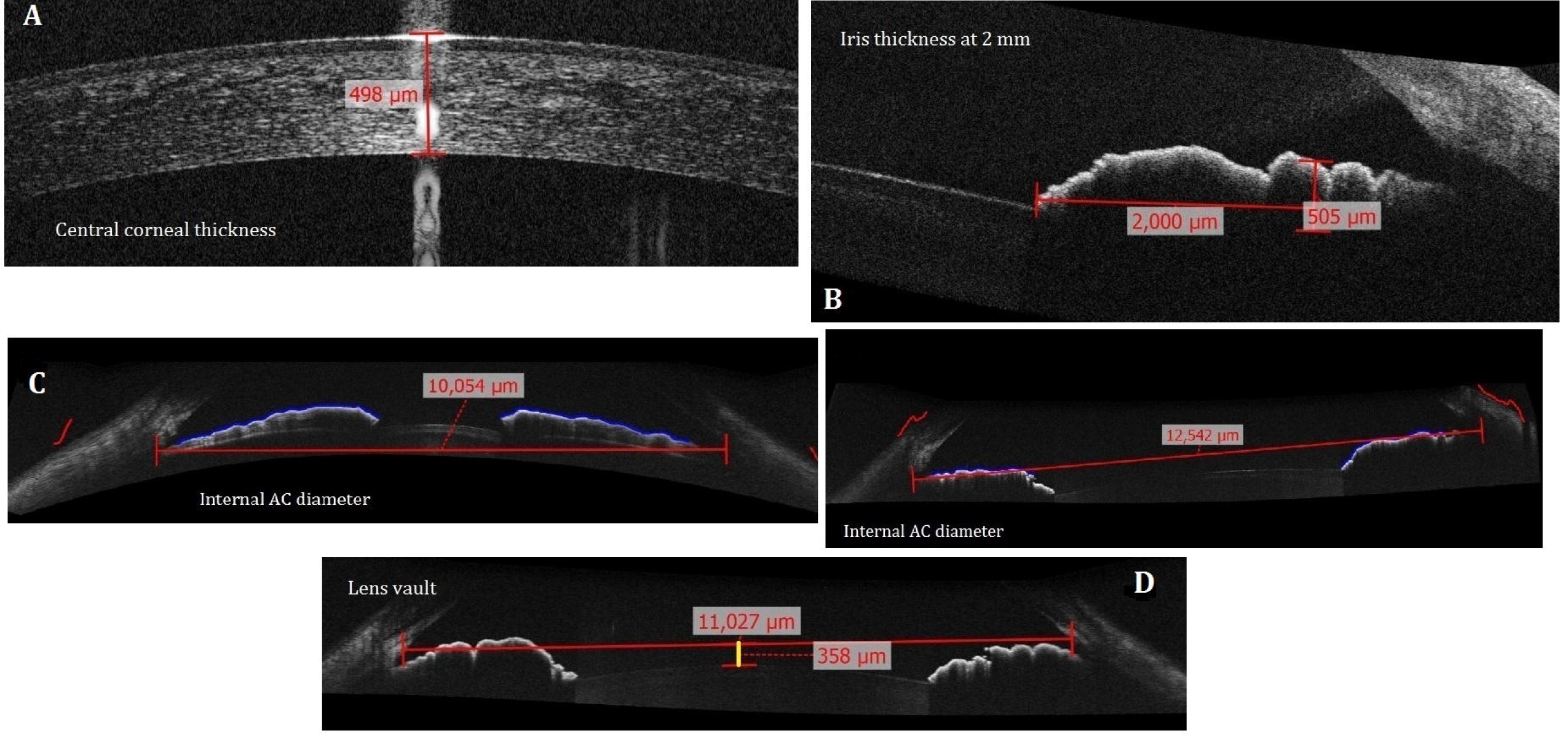
A: Central corneal thickness B: Iris thickness at 2 mm C: Internal AC diameter in shallow AC and deep AC D: Lens vault estimation

#### 2.1.2. Iris thickness (µm)

was calculated at 2 mm from the margin of the pupil, both nasally and temporally at 3 and 9 o’clock. [Figure 2B]

#### 2.1.3. Internal AC diameter (µm)

was manually calculated from the wide scan image by measuring the distance between the two angle recesses. The horizontal distance was measured for this purpose. [Figure 2C]

#### 2.1.4. Lens Vault (LV) distance (µm)

is the perpendicular distance between the anterior pole of the crystalline lens and the horizontal line drawn between the two scleral spurs, on horizontal AS-OCT scans. For manual measurements, the ‘*line measurement tool’* was utilised, which is built in the software. [Figure 2D]

#### 2.1.5. The *anterior chamber (AC) angle*

is the junction between the root of the iris and the cornea. The scleral spur is the most vital anatomical landmark in angle evaluation, which connects the posterior corneal curvature to the scleral curvature. The ***AC angle*** was determined by the ‘*angle measurement tool’* of the OCT program by manually locating the apex and tracing the angle between the iris and posterior cornea. The measurement is expressed in degrees.

#### 2.1.6. The *angle opening distance (AOD) (mm)*

is the distance between the corneal endothelium and the anterior iris surface along a perpendicular line drawn across the trabecular meshwork at 500 μm and 750 μm anterior to the scleral spur [7, 8]. The ***AOD 500*** and ***AOD 750*** are calculated by the ‘*AOD tool’* of the OCT program, placing the tool at the scleral spur and then corneal endothelium and iris, and the program calculates the measurements automatically.

#### 2.1.7. The *trabecular-iris space area (TISA)*(mm^2^)

is the trapezoidal area with boundaries formed anteriorly by the AOD (500 or 750), posteriorly by the inner scleral wall, inner superiorly by the corneoscleral wall, and inferiorly by the iris surface. Its posterior border is demarcated by a line drawn from the scleral spur to the iris perpendicular to the inner scleral wall [8]. ***TISA 500*** and ***TISA 750*** are measured with the AOD tool as well.

#### 2.1.8. The *TIA (trabecular iris angle)*

in degrees is also measured with this tool. TIA 500 is defined as an angle with its apex in the iris recess and its arms passing through perpendicular points on the trabecular meshwork 500 μm from the scleral spur and the iris [9]. [Figure 3]

**Figure 3:**
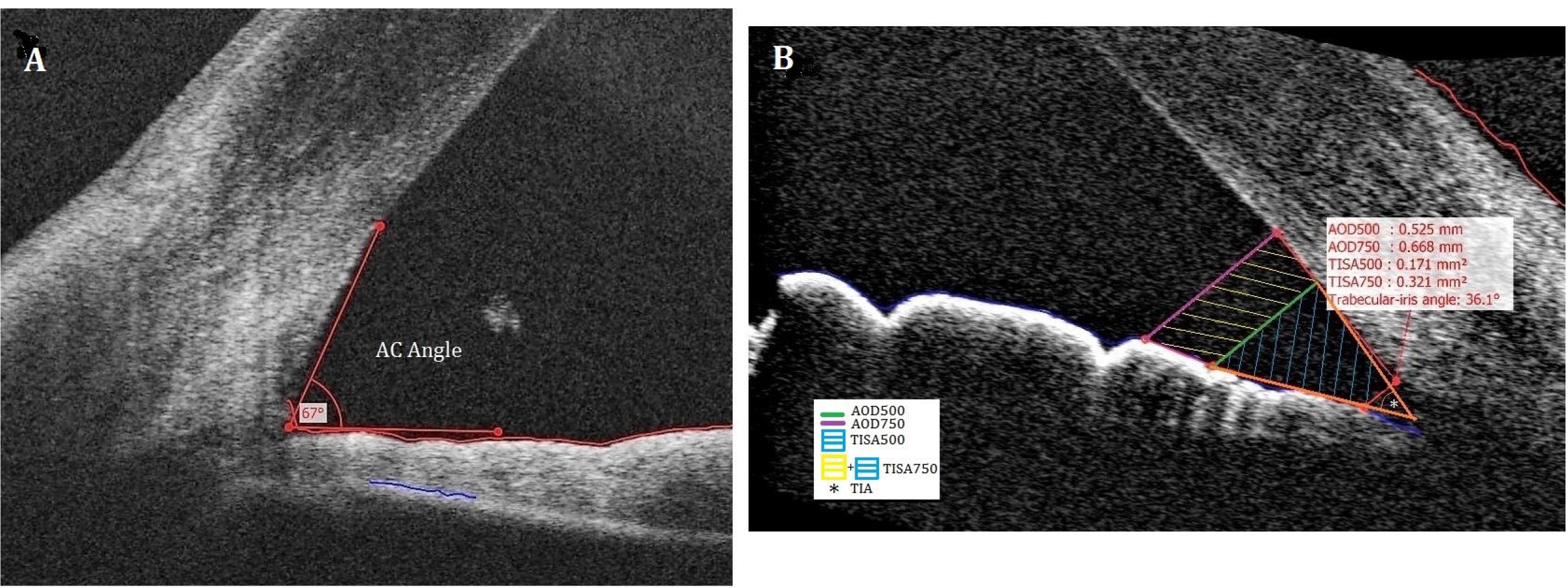
A. *Anterior chamber (AC) angle measurement B. AOD500, AOD750, TISA500, TISA750 and TIA measurement with the AOD tool [AOD: angle opening distance TISA: trabecular-iris space area TIA: trabecular iris area]*

All measurements were done for both the eyes separately. The 3 and 9 o’clock positions were used for analysis.

The data was tabulated and analyzed via SPSS version 20. Frequencies and percentages were calculated for age, gender, IOP, axial length, CDR, spherical equivalent, CCT, internal AC diameter, AC angle, AOD, TISA, TIA, iris thicknesses and lens vault. The independent t-test was used to assess variable difference between genders. The paired samples t-tests were used to analyze the differences between the nasal and temporal parameters. Pearson’s correlation coefficient was used to assess relationship between all the variables.

## 3. RESULTS

Results were analyzed via SPSS version 20. Mean age of the patients was 11.38±3.18 years [range 3-16] with a female predominance; 29 (53.7%) patients and 58 eyes. Mean IOP was 13.63±2.90 mmHg [5-21], mean axial length was 22.82±0.89 mm, mean spherical equivalent was -0.14±1.28 D [-3.63-+3.75], mean CCT was 532.6±46.09mm [434-721] and mean CDR was 0.31±0.19. The mean internal AC diameter was 11609.15±2354 µm. The paired samples t-tests were used to analyze the differences between the nasal and temporal parameters. The nasal AC angle was 53.54±11.82° and significantly wider than the temporal AC angle; 50.37±10.26° [p=0.008]. The nasal AOD500 was 0.9±0.33 mm and not significantly greater than the temporal AOD500, 0.85±0.28 mm [p=0.09]. The nasal AOD750 was 1.21±0.45 mm, significantly greater than the temporal AOD750, 1.06±0.32 mm [p=0.001]. The nasal TIA was 41.75° and not significantly greater than the temporal TIA, 40.24° [p=0.246]. The nasal TISA500 was 0.33±0.15 mm^2^ and not significantly wider than the temporal TISA500, 0.31±0.10 mm^2^ [p=0.056]. The nasal TISA750 was 0. 59±0.22 mm^2^ and not significantly wider than the temporal TISA750, 0.56±0.21 mm^2^ [p=0.14]. The nasal iris thickness at 2 mm from pupil was 483.54±129.56 µm, and not significantly different from the temporal iris thickness, 505.8±138.85 µm [p=0.08]. The mean lens vault was -519.58±359µm [+331 to -1584]. [Table 1]

**Table 1:** Anterior segment characteristics of healthy Pakistani children

|  | *Mean ± Std. Deviation* | *Minimum* | *Maximum* |
| --- | --- | --- | --- |
| Age | 11.38 ± 3.18 | 3 | 16 |
| IOP mmHg | 13.63 ± 2.90 | 5 | 21 |
| CCT $\mu\text{m}$ | 532.6 ± 46.09 | 434 | 721 |
| CDR | 0.31 ± 0.19 | 0.00 | 0.75 |
| Spherical Equivalent D | -0.14 ± 1.28 | -3.63 | +3.75 |
| Axial Length mm | 22.82 ± 0.89 | 20.91 | 25.15 |
| Internal AC diameter $\mu\text{m}$ | 11609.15 ± 2354 | 11.9 | 13476 |
| AC angle Nasal ° | 53.54 ± 11.82 | 31 | 85 |
| AC angle Temporal ° | 50.37±10.26 | 26 | 81 |
| AOD500 Nasal mm | 0.90 ± 0.33 | 0.35 | 2.13 |
| AOD750 Nasal mm | 1.21 ± 0.45 | 0.27 | 2.80 |
| AOD500 Temporal mm | 0.85 ± 0.28 | 0.33 | 1.99 |
| AOD750 Temporal mm | 1.06 ± 0.32 | 0.48 | 2.38 |
| TIA Nasal ° | 41.75 ± 11.49 | 8.90 | 61.9 |
| TIA Temporal ° | 40.24 ± 10.52 | 16.7 | 69.2 |
| TISA500 Nasal mm<sup>2</sup> | 0.33 ± 0.15 | 0.12 | 0.99 |
| TISA750 Nasal mm<sup>2</sup> | 0.59 ± 0.22 | 0.24 | 1.52 |
| TISA500 Temporal mm<sup>2</sup> | 0.31 ± 0.10 | 0.10 | 0.84 |
| TISA750 Temporal mm<sup>2</sup> | 0.56 ± 0.21 | 0.22 | 1.62 |
| Iris thickness at 2mm Nasal $\mu\text{m}$ | 483.54 ± 129.56 | 215 | 962 |
| Iris thickness at 2mm Temporal $\mu\text{m}$ | 505.8 ± 138.85 | 242 | 879 |
| Lens Vault $\mu\text{m}$ | -519.58 ± 359 | +331 | -1584 |

AC: anterior chamber AOD: angle opening distance TIA: Trabecular iris angle TISA: Trabecular iris space area

Pearson’s correlation coefficient was used to assess relationship between all the variables. There was no correlation of any variable with age except temporal iris thickness [p=0.007] and axial length [p=0.01]. Analysis of variance (ANOVA) was used to assess gender differences. The IOP was significantly lower in males [p=0.001]. The temporal AC angle was wider in females [p=0.038]. [Table 2]

**Table 2:**
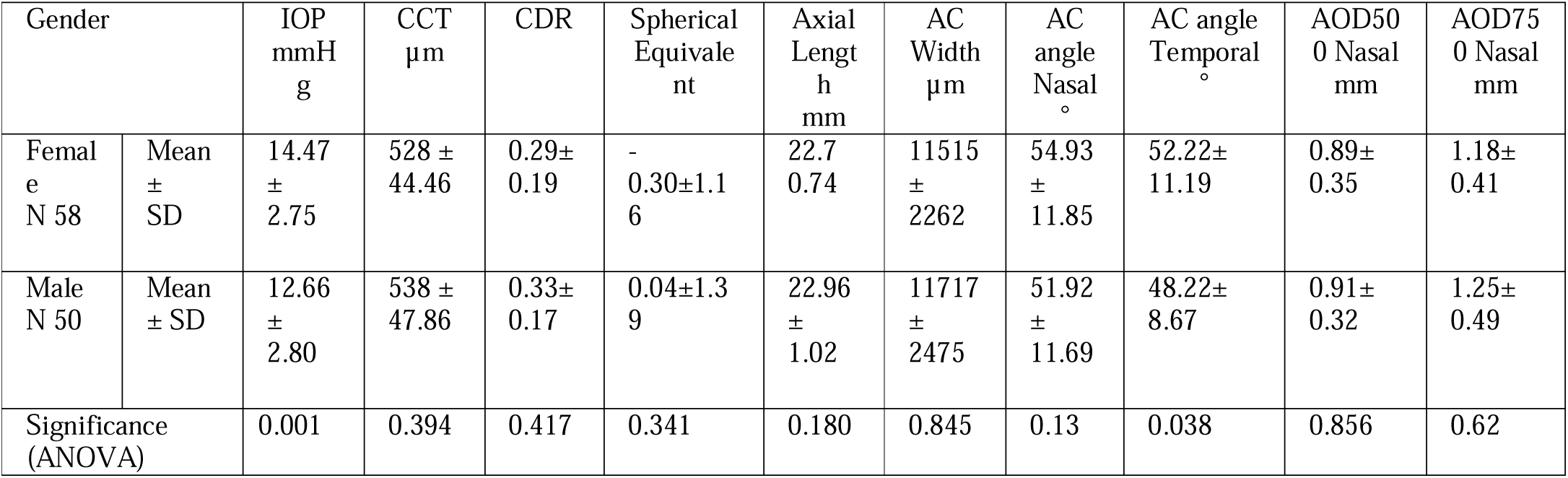
Gender differences amongst anterior segment parameters

A longer axial length was associated with a negative lens vault [p=0.001]. Wider AC width was associated with IOP [p=0.01], nasal AC angle [p=0.04], and temporal AC angle [p=0.000].

## 4. DISCUSSION

AS-OCT is gaining importance in anterior segment evaluation and quantification and has immense utility. It is also useful in longitudinal assessment of anatomy and developmental changes in children. It is a valuable tool for diagnostics, therapeutics and prognostics pertaining to the anterior segment [10, 11].

Wang et al in their Pentacam Scheimpflug camera study on Chinese school children found the mean AC angle to be 37.95±7.9°, whereas in our population, the nasal AC angle was 53.54±11.82° and the temporal AC angle was 50.37±10.26°, which is wider than the Chinese children [12]. Wang found higher values in boys as compared to girls, whereas in our study, only the temporal AC angle is significantly wider in females and IOP is significantly lower in males. However, our study also shows that males have thicker corneas [538:528 µm], higher CDR [0.33:0.29], longer axial lengths[22.9:22.7], wider anterior chambers[11717:11515 µm], wider nasal AOD500[0.91:0.89 mm] and AOD750[1.25:1.18mm], wider nasal TIA [43.2:40.5°], wider nasal TISA750 [0.60:0.59mm^2^]and temporal TISA750 [0.59:0.53mm^2^], and a greater negative lens vault [-547:-495µm]compared to females; but not significantly so. Females in our study have wider AC angles, both nasally [54.9:51.9°] and temporally [52.2:48.2°] compared to males, wider temporal TIA [40.33:40.15°], a wider TISA500 nasally[0.34:0.33mm^2^] and thicker irides at 2mm both nasally[488:479µm] and temporally [515:495 µm] compared to males. [Table 2 and 3]

**Table 3:** Gender differences amongst angle parameters, iris thickness and lens vault

| Gender |  | TIA<br>Nasal<br>° | TIA<br>Temporal<br>° | TISA<br>500<br>Nasal<br>mm<sup>2</sup> | TISA<br>750<br>Nasal<br>mm<sup>2</sup> | TISA<br>500<br>Temporal<br>mm<sup>2</sup> | TISA<br>750<br>Temporal<br>mm<sup>2</sup> | Iris<br>thickne<br>ss<br>at 2mm<br>Nasal | Iris<br>thickness<br>at 2mm<br>Temporal | Lens<br>Vault<br>µm |
| --- | --- | --- | --- | --- | --- | --- | --- | --- | --- | --- |
| Femal<br>e<br>N 58 | Mean<br>±<br>SD | 40.5 ±<br>11.74 | 40.33±<br>11.21 | 0.34±<br>0.16 | 0.59±<br>0.23 | 0.29±<br>0.08 | 0.53±<br>0.15 | 488±<br>123 | 515±<br>144 | -495±<br>313 |
| Male<br>N 50 | Mean<br>±<br>SD | 43.2 ±<br>11.14 | 40.15±<br>9.77 | 0.33±<br>0.12 | 0.60±<br>0.21 | 0.33±<br>0.12 | 0.59±<br>0.25 | 479±<br>137 | 495±<br>134 | -547±<br>408 |
| Significance<br>(ANOVA) |  | 0.438 | 0.821 | 0.717 | 0.867 | 0.075 | 0.053 | 0.492 | 0.409 | 0.453 |

Jin et al similarly studied anterior segment parameters in normal Chinese children using Fourier domain OCT (FD-OCT), and found no significant differences in angle parameters between sexes, but found an association of age with increasing AC width [13]. In a study on iris thickness by Nakakkura et al, the average iris thickness (temporal and nasal) at 1 and 2 mm were 432 ± 0.06[302-569 µm] and 337 ± 0.04 [229-414 µm], respectively in Japanese children [14]. We studied iris thickness at 2mm and found it to be 483.54 ± 129 µm[215-962] nasally and 505.8 ± 139 µm [242-879] temporally, in our population. Our children have thicker irides than Japanese children.

Shimizu et al in their study comparing children and adults found that children have thicker CCT (560μm), deeper anterior chamber (3.05mm) and larger AOD (0.56mm) as compared to adults (all P < 0.01). The lens vault was smaller in the children than in the adults (0.04mm) [15]. Qiu et al have used OCT to evaluate emmetropic and myopic children and found the anterior segments to be elongated in the latter, there being no significant difference of CCT between children with myopia (551:548 µm) versus children with emmetropia; the angle width was also similar in both groups [16]. The AOD500 of inferotemporal quadrant was widest in both groups; the average AOD500 being wider in myopic group when compared with emmetropic group. Fernández-Vigo et al studied trabecular meshwork and TIA in healthy white children using FD-OCT, with mean TIA, being 43.1±10.0° [17]. They also found nasal ACA width to be wider 45.9±9.1°; than in the temporal quadrant in their study; 44.3±10.1° [18].

The importance of assessing these parameters in normal children is to distinguish them from pathological eyes especially in pediatric glaucomas, but also other anterior segment disorders. Bradfield et al used the AS-OCT modality to investigate normal pediatric eyes versus glaucomatous eyes and noted absence of Schlemm’s canal (SC) in the glaucoma affected eyes only, and abnormal tissue over the angle and the SC [19]. Pilat et al found thinner collarettes and flat rides in primary congenital glaucoma versus healthy controls using a hand-held OCT device [20]. Chen et al have studied anterior segment parameters in pediatric eyes which developed secondary glaucoma after cataract surgery using ultrasound biomicroscopy to understand predisposing factors [21]. Monsálvez-Romín et al studied adult eyes during accommodation using Visante OCT [22]. Hashemi et al used the Lenstar biograph for anterior chamber assessment in children; mean axial length was 23.13 mm (22.93–23.33), anterior chamber depth (ACD) was 3.01 mm (2.96–3.06), lens thickness (LT) was 3.58 mm (3.55–3.61), central corneal thickness (CCT) was 549.33 µm (546–552), corneal radius (CR) was 7.77 mm (7.74–7.81), corneal diameter (CD) was 12.34 mm (12.31–12.38) and pupil diameter (PD) was 4.97 (4.91– 5.03). Mean AL (axial length), ACD (AC depth), CD (corneal diameter) and CR (corneal radius) were found to be significantly higher in boys, whereas the mean LT (lens thickness) was significantly higher in girls. The AL and ACD increased with age, while LT decreased significantly so [23]. Edawaji et alhave used the hand held AS-OCT to effectively study the anterior chamber development in newborns [24].

Strengths of our study are that it is a very comprehensive and detailed study of anterior segment characteristics in the normal pediatric population. It is also the first of its kind in our country.

Limitations of our study are that it is a single center study with a moderately small sample size. Also, ACD, ACV (AC volume), lens thickness, trabecular meshwork and ciliary body have not been analyzed. This is because currently our OCT machine does not have a biometry attachment. Swept source OCT is more useful for deeper structure assessment, but may have limitations also, especially in viewing ciliary body and structures in the presence of opaque media [25].

Future work includes assessment of angle parameters in pediatric eyes with ocular diseases, especially glaucomas, both primary and secondary, as well as other diseases of anterior segment, and congenital anomalies. Comparison of normative data with diseased eye data is likely to yield useful differences. There is more to learn.

## 5. CONCLUSIONS

**1**. Our study data depicts the normal anterior segment parameters in healthy children. This will be useful in diagnosing and managing ocular pediatric pathology.

## Data Availability

The data for this study is available from Open Science Framework with the DOI 10.17605/OSF.IO/BRS2H and link: https://osf.io/brs2h/?view_only=eb70e5865b6048969a29e61d21dcf7d3

## Author contribution

Conceptualization; Data curation; Formal analysis; Investigation; Methodology; Project administration; Resources; Software; Supervision; Validation; Visualization; Roles/Writing - original draft; Writing - review & editing

